# Comparison of immunogenicity of adjuvanted and high dose influenza vaccination in long-term care facility residents

**DOI:** 10.1101/2024.10.14.24315459

**Authors:** Elise M. Didion, Joseph D. Kass, Dennis J. Wilk, Emily Buss, Sarah-Michelle Frischmann, Sabina Rubeck, Richard Banks, Brigid M. Wilson, Stefan Gravenstein, David H. Canaday

**Affiliations:** Case Western Reserve University School of Medicine, Cleveland, OH; Geriatric Research, Education and Clinical Center at Louis Stokes Cleveland VA Medical Center, Cleveland, OH; Department of Molecular Biology and Microbiology, School of Medicine, Case Western Reserve University; Department of Health Services, Policy, and Practice, Brown University School of Public Health, Providence, RI; Center on Innovation in Transformative Health Systems Research to Improve Veteran Equity and Independence, Providence Veterans Administration Medical Center, Providence, RI; Division of Geriatrics and Palliative Medicine, Alpert Medical School of Brown University, Providence, RI

**Keywords:** Adjuvanted vaccine, High-Dose vaccine, Influenza vaccine, Long-term care, Aged, Immunogenicity

## Abstract

**Background:** The CDC recommends the more immunogenic adjuvanted and high-dose flu vaccines over standard-dose, non-adjuvanted vaccines for individuals above 65 years old. The current study compares adjuvanted trivalent inactivated flu vaccine (aTIV, FLUAD) versus high-dose flu vaccine (HD-IIV3, FLUZONE HD) to determine if they met non-inferiority standards for older long-term care facility (LTCF) residents.

**Methods:** We collected blood from long-term care facility residents participating in a randomized 1:1 active control trial comparing MF59C.1 adjuvanted trivalent inactivated flu vaccine, aTIV versus HD-IIV3 over the course of two flu seasons, 2018-2019 and 2019-2020 (Trial, NCT03694808). We assessed humoral immunity at set time points via hemagglutinin inhibition assays (HAI) and anti-neuraminidase (enzyme-linked lectin assays (ELLA)). The recombinant influenza vaccine (RIV, Flublok) was assessed similarly in year two for a small number of participants who were carried over from year 1 (n=32).

**Results:** We enrolled 387 volunteers and administered either aTIV (n=194), HD-IIV3 (n=193) over the course of the 2018-2019 and 2019-2020 flu seasons. Among those enrolled and randomized in year one, a subset were administered RIV and studied in year two (n = 32). At 28 days post-vaccination, aTIV exhibited non-inferiority to HD-IIV3 for HAI for both H1N1 and H3N2 strains (GMT ratios (95% CI) for HD-IIV3/aTIV of 1.03(0.76, 1.4) and 1.04(0.73, 1.48), respectively; both 95% CI upper bounds < 1.5 to meet non-inferiority criteria) but not for Influenza B (GMT ratio (95% CI) = 1.21 (0.91, 1.61)). Non-inferiority criteria for HAI seroconversion were not met for any of the three strains. Applying the same non-inferiority criteria to neuraminidase inhibition (NI), both day 28 titer and seroconversion in aTIV were non-inferior to HD-IIV3 for H1N1 and H3N2 strains.

**Conclusions:** Both aTIV and HD-IIV3 elicited similar immune responses with robust antibody responses. For the primary outcome, aTIV is non-inferior to HD-IIV3 for HAI titer of H1N1 and H3N2 but failed to meet non-inferiority criteria for Influenza B and seroconversion for all assessed strains. For the secondary outcome, aTIV was non-inferior to HD-IIV3 for both titer and seroconversion of anti-neuraminidase for both H1N1 and H3N2.

## Background

Influenza continues to cause significant morbidity and mortality globally, particularly affecting adults over the age of 65 and those residing in long-term care facilities (LTCFs) Increased frailty, comorbidities, and communal living arrangements make this population especially vulnerable^1^. Standard-dose influenza vaccines have reduced effectiveness in older adults, prompting the development of formulations that elicit stronger immune responses. The Centers for Disease Control and Prevention (CDC) currently recommend the preferential use of adjuvanted (aTIV), high-dose (HD-IIV3) and recombinant (RIV) influenza vaccines for individuals over 65 years of age ^2^.

The CDC approved enhanced vaccines, such as Fluzone High-Dose (HD-IIV3; Sanofi Pasteur Inc.) in 2009, which contains four times the hemagglutinin (HA) antigen content of standard-dose vaccines. HD-IIV3 vaccine has demonstrated a greater effectiveness at preventing influenza in aging populations including LTCF residents compared to standard-dose vaccines ^3, 4^. The CDC also approved an influenza vaccine enhanced with the MF59 adjuvant, Fluad (aTIV; Seqirus USA Inc.), for older adults in 2016. Various studies have demonstrated the efficacy of adjuvants in enhancing immune responses, and greater effectiveness even in elderly populations ^5, 6^.

This study aims to compare the immunogenicity of the adjuvanted aTIV vaccine with the high-dose HD-IIV3 vaccine to determine whether aTIV is non-inferior in older adults residing in LTCFs. We conducted a randomized, controlled trial over two influenza seasons (2018-2019 and 2019-2020) to assess the humoral immune responses elicited by these vaccines using hemagglutination inhibition (HAI) and enzyme-linked lectin assays (ELLA assay). Our primary outcome measures included geometric mean titer (GMT) ratios for H1N1, H3N2, and influenza B strains at 28 days post-vaccination. Secondary outcomes included seroconversion rates and neuraminidase (NA) inhibition (NI) titers. Given the variability in vaccine performance across different influenza seasons, this study provides valuable insights into the comparative effectiveness of these two enhanced vaccines in a highly susceptible population.

With both seasons combined we found that aTIV met HAI GMT non-inferiority standards for H1N1 and H3N2 but not B, however, aTIV failed to meet non-inferiority standards for HAI seroconversion. For neuraminidase, aTIV is non-inferior to HD-IIV3 for both GMT and seroconversion. Similar to other studies ^7^, results differed between flu seasons suggesting that the ability of these two vaccines to elicit immune responses may not greatly differ.

## Methods

### Trial design and oversight

We conducted a phase 4 randomized active-control trial comparing the aTIV vaccine to the HD-IIV3 vaccine. The trial included participants aged 65 years and older residing in long-term care facilities across Ohio. The study spanned two influenza seasons, from September 23, 2018, to December 30, 2020. The University of Hospitals of Cleveland and the Cleveland VA Institutional Review Boards (IRB) approved the study which complied with international standards and was registered on ClinicalTrials.gov on 13 October 2018 (Identifier: NCT03694808). We obtained written informed consent from all participants or their legally authorized representatives.

### Participants and group assignments

Participants were long-term care residents aged 65 and older who did not have recent acute illnesses, were not using immunomodulatory agents, and had no recent history of cancer requiring treatment, certain circulatory system issues, allergies or reactions to any component of the influenza vaccine, or a history of Guillain-Barré syndrome following influenza vaccination.

We randomized participants in a 1:1 ratio to receive either the aTIV or HD-IIV3 vaccine. Blood samples were collected at baseline (D0, -7-0 days), day 28 (D28, 24-29 days), and for the 2018-2019 season, day 180 (D180, 192-215 days). Day 180 samples were not obtained for 2019-2020 due to the start of the SARS-CoV-2 pandemic. Samples were processed using standard methods and stored at -80°C until analysis.

### Vaccines

HD-IIV3 is derived from inactivated influenza virus amplified in chicken eggs and concentrated to present fourfold more HA per strain (60 mcg HA per strain per dose) than the standard dose vaccine. aTIV is also generated from chicken egg grown influenza virus with the addition of MF59C.1 adjuvant (MF59®), a squalene-based oil-in-water emulsion at 15 mcg HA per strain per dose (Seqirus USA Inc., NJ USA). RIV comes from a baculovirus vector driven recombinant process generating the HA subunit, and includes the purified HA at a 45mcg/ml amount per strain per dose. The 2019-2020 RIV formulation was quadrivalent and contained 2 B strains while aTIV and HDIIV3 are both trivalent formulations.

### Assays

#### Hemagglutinin Inhibition Assays (HAI)

HAI detects the presence of HA-specific antibodies in donor sera following previously established protocols (Klimov, 2012) using turkey red blood cells (Lampire Biological Laboratories). We acquired strain-matched virus for A/H1N1 (A/Michigan/45/2015, FR1483), A/H3N2 (A/Singapore/02/2018, FR-1590), and influenza B (B/Colorado/01/2018, FR-1588) from International Reagent Resource (IRR) for the 2018-2019 flu season. For the 2019-2020 flu season, A/H1N1 (A/Brisbane/02/2018, FR1665) and A/H3N2 (A/Kansas//14/2017, FR1666) from Scott Hensley, University of Pennsylvania, and influenza B (B/Colorado/06/Victoria, FR1667) were utilized. Immunogenicity was measured by geometric mean titers (GMT), seroprotection, and seroconversion rates.

#### Neuraminidase Inhibition (NI) measuring anti-neuraminidase titers

We used standard methods of the anti-NA enzyme-linked lectin assays (ELLA) to determine neuraminidase inhibition activity ^8^. Briefly, Immulon 4 HBX plates (ThermoFisher Scientific, Waltham, MA) were coated with fetuin (Sigma F3385 at 25 ug/ml) and incubated overnight at 4°C. Sera was heat treated at 56°C and serially diluted across the plate prior to the addition of strain-matched enzymatically active recombinant NA (SinoBiological, Chesterbrook, PA) for each season. Plates were incubated overnight at 37°C, washed, and peanut agglutinin horseradish peroxidase conjugate (SIgma L7759) was added for 2 hours at room temperature (RT). Plates were visualized with a citrate buffer and an o-phenylenediamine dihydrochloride tablet (Sigma; P8287). After a 10-minute incubation at RT, the reaction was stopped with 0.5M sulfuric acid and read at 490nm.

### Analysis

All analyses were performed in R Version 4.2.2. Seroconversion was defined as a fourfold or greater increase in HAI titer at day 28, and seroprotection was defined as an HAI titer ≥40. The GMT at day 28 was used to assess non-inferiority, considering the ratio and 95% confidence interval of the HD-IIV3 GMT to that of aTIV. For aTIV to be considered non-inferior to HD-IIV3 by FDA guidelines, the upper bound of this two-sided 95% CI could not exceed 1.5.

For seroconversion rates, non-inferiority of aTIV to HD-IIV3 was achieved if the upper bound of the two-sided 95% CI for the difference in rates (HD-IIV3 - aTIV) did not exceed 10 percentage points. We compared baseline levels of HAI and NI titers between aTIV and HD-IIV3 vaccines using standard mean differences (SMD) to assess the balance achieved by the randomization, with absolute values <0.1 considered well-balanced and >0.25 considered strongly unbalanced ^9^. Additionally, we summarized titers observed at day 180 where available in the year one cohort; and the day 0 and day 28 RIV, aTIV, and HD-IIV3 titers in the year two cohort in which the third vaccine RIV was administered to a subset as well.

## Results

### Study participants

We screened 478 participants for inclusion in the study and enrolled and randomized 387 to receive either aTIV or HD-IIV3. An additional 32 participants from the first year of the study also participated in the second year, receiving RIV as their administered vaccine. Common reasons for exclusion from the study were withdrawn consent or unsuccessful/missed blood draws at the designated time points. The participants ranged in age from 65 to 100 years at the time of enrollment, with an approximately equal distribution of male and female participants and 20% of participants identifying as non-White. Similar distributions of age, gender, and race were observed between the aTIV and HD-IIV3 groups (Table 1). Baseline levels of both HAI and NI titers were also found to be balanced between the vaccine groups (Supplemental Table 1), with absolute standardized mean differences not exceeding 0.1 for the two years combined nor 0.25 for either study year.

**Table 1.**
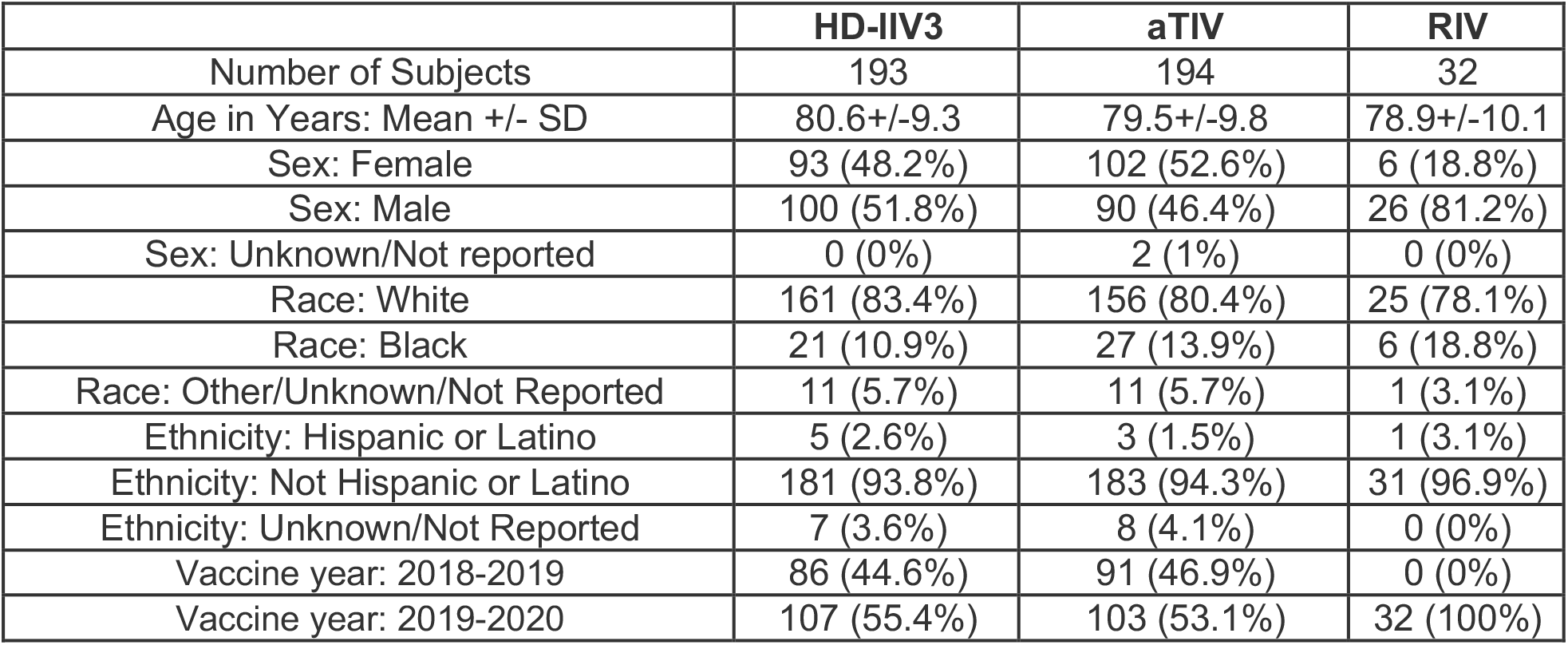
Demographics of subjects by vaccine.

### Primary Outcome

When combining data from both flu seasons (Figure 1), the HAI GMTs for H1N1, H3N2, and B strains for subjects administered aTIV were 117.8, 222.5, and 91.4, respectively. For subjects administered HD-IIV3, these GMTs were 121.7, 230.5, and 110.7. Based on the 95% confidence interval about the ratios of the HD-IIV3 GMT to the aTIV GMT, aTIV met non-inferiority criteria for HAI GMT ratios at 28 days post-vaccination for both H1N1 and H3N2 but not for the influenza B strain (Table 2).

**Table 2.** Statistical summary of HAI GMT and seroconversion rate at Day 28 for aTIV and HD-IIV3. HD-IIV3 shows GMT NI for H1N1 and H3N2 (bold & italicized) where upper bound of 95% confidence interval about the GMT ratio of HD-IIV3 to aTIV is < 1.5. This statistic exceeds 1.5 for B strain, and thus does not demonstrate non-inferiority. aTIV seroconversion fails to meet non-inferior criteria to HD-IIV3 for H1N1, H3N2 and B as all upper bounds of seroconversion differences 95% CI exceed 0.1.

| Strain | HD-IIV3 |  |  | aTIV |  |  | HD-IIV3 vs. aTIV comparisons |  |
| --- | --- | --- | --- | --- | --- | --- | --- | --- |
|  | N subjects | Day 28 GMT | N (%) Serocon-version | N subjects | Day 28 GMT | N (%) Serocon-version | GMT Ratio (95% CI) | Seroconversion difference (95% CI) |
| B | 193 | 110.7 | 83 (43%) | 194 | 91.4 | 69 (36%) | 1.21 (0.91,1.61) | 0.074 (-0.028, 0.177) |
| H1N1 | 190 | 121.7 | 83 (44%) | 192 | 117.8 | 76 (40%) | **1.03 (0.76,1.4)** | 0.041 (-0.063, 0.145) |
| H3N2 | 193 | 230.5 | 124 (64%) | 194 | 222.5 | 105 (54%) | **1.04 (0.73,1.48)** | 0.101 (-0.001, 0.204) |

**Figure 1.**
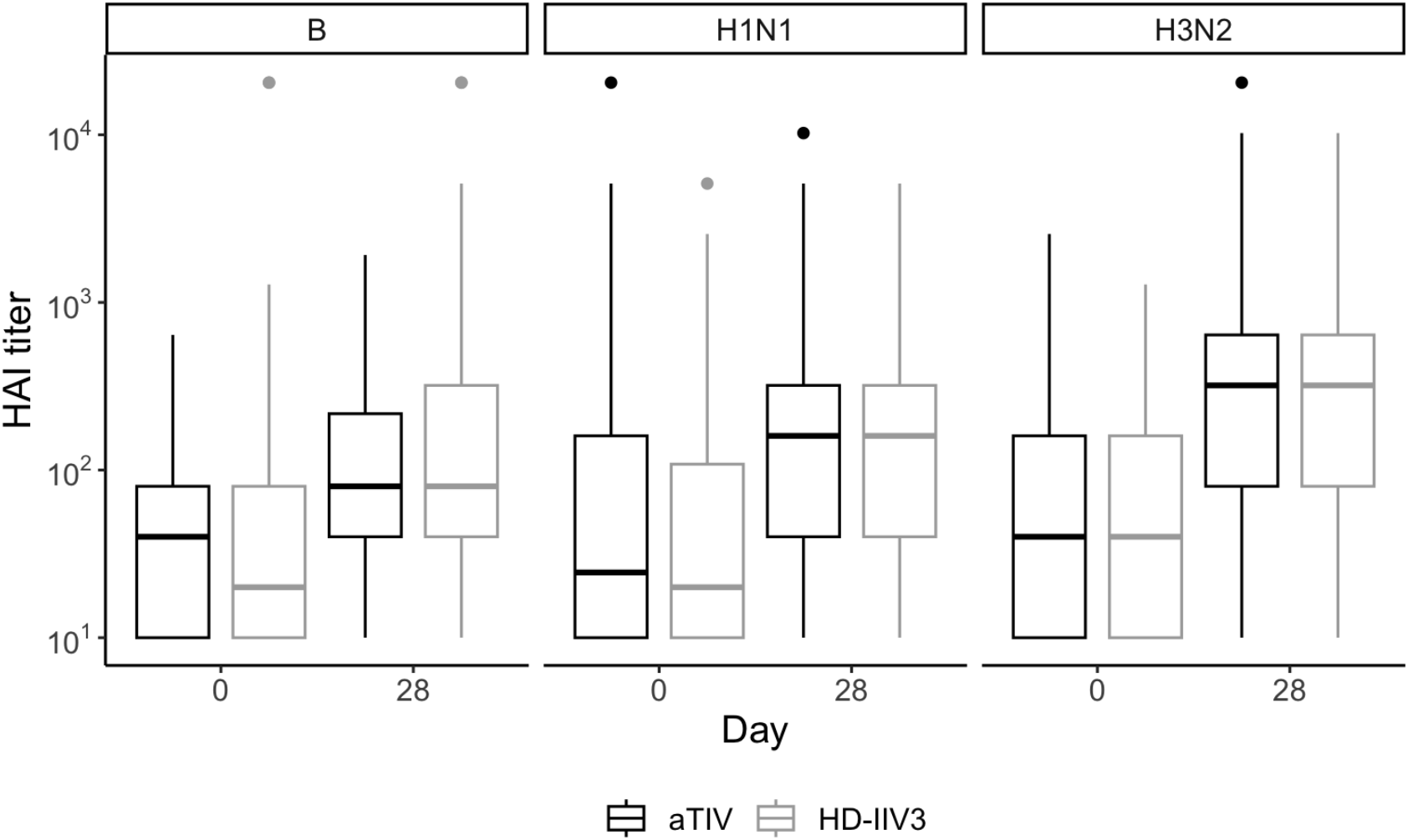
Observed distributions of HAI titer for aTIV (black) and HD-IIV3 (gray), by day and strain. For each box, the center line indicates the median and the bottom and top of the box indicate the first and third quartile, respectively. The lower and upper whiskers extend from the first and third quartile lines, respectively, to the smallest and largest values no more than 1.5 times the interquartile range (height of box) away from the first and third quartile values.

**Figure 2.**
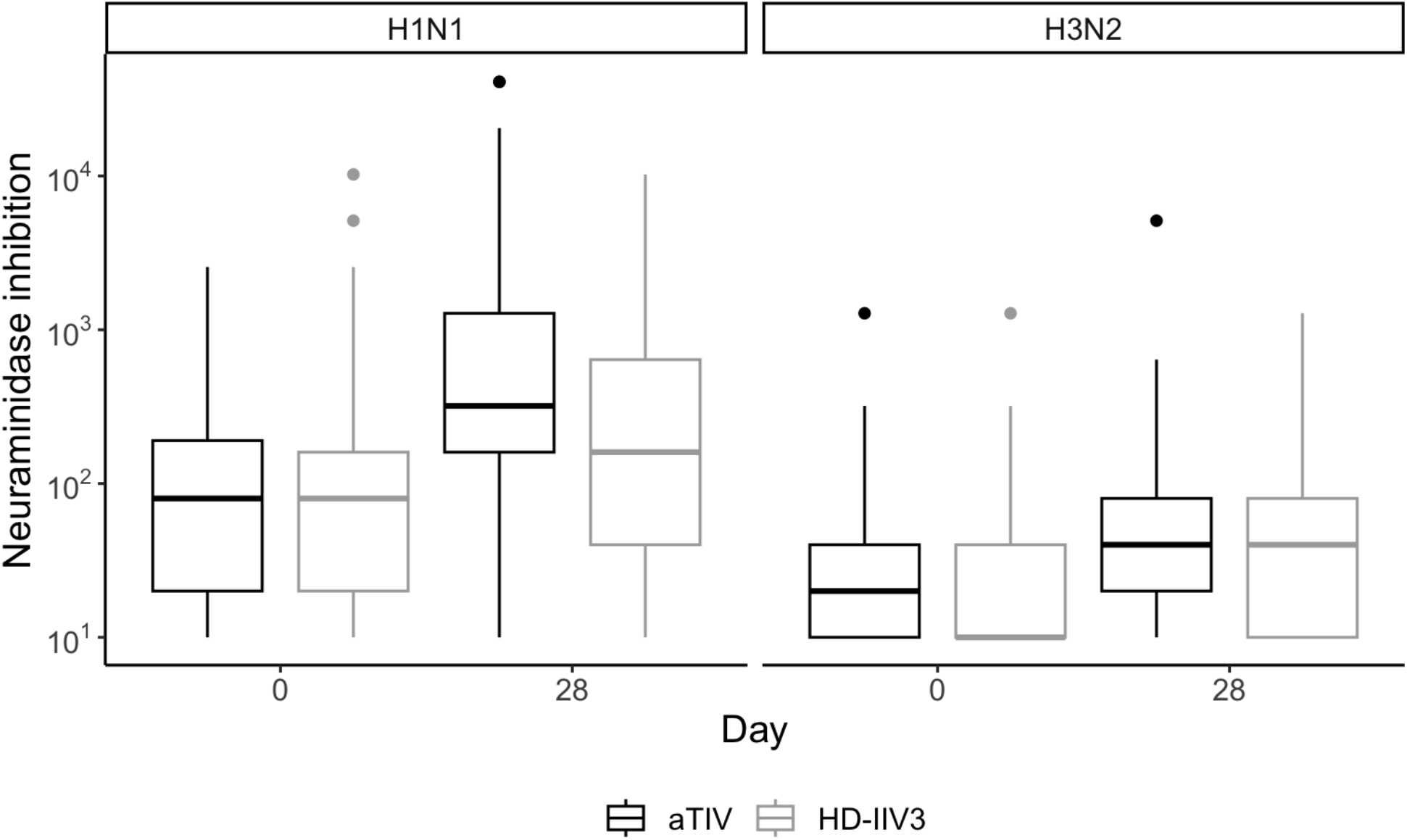
Observed distributions of NI titers for aTIV (black) and HD-IIV3 (gray), by day and strain. For each box, the center line indicates the median and the bottom and top of the box indicate the first and third quartile, respectively. The lower and upper whiskers extend from the first and third quartile lines, respectively, to the smallest and largest values no more than 1.5 times the interquartile range (height of box) away from the first and third quartile values.

Combining data from both flu seasons, seroconversion rates at day 28 for participants who received aTIV were 39.6% for H1N1, 54.1% for H3N2, and 35.6% for the B strain. In participants who received HD-IIV3, the seroconversion rates were 43.7% for H1N1, 64.2% for H3N2, and 43.0% for the B strain (Table 3), and the non-inferiority criteria was not met for any strain.

**Table 3.** Statistical summary of NI GMT and seroconversion for aTIV and HD-IIV3. For both measures, aTIV shows non-inferiority to HD-IIV3 for both H1N1 and H3N2 (bold and italicized), with the upper limit of the HD-IIV3 to aTIV GMT ratio 95% confidence interval not exceeding 1.4 and the upper limit of the seroconversion difference 95% confidence interval not exceeding 0.1.

|  | HD-IIV3 |  |  | aTIV |  |  | HD-IIV3 vs. aTIV comparisons |  |
| --- | --- | --- | --- | --- | --- | --- | --- | --- |
| Strain | N subjects | Day 28 GMT | N (%) Seroconversion | N subjects | Day 28 GMT | N (%) Seroconversion | GMT Ratio (95% CI) | Seroconversion difference (95% CI) |
| H1N1 | 189 | 159.4 | 54 (29%) | 192 | 351.5 | 119 (62%) | ***0.45 (0.33,0.63)*** | ***-0.334 (-0.433, -0.235)*** |
| H3N2 | 192 | 34.4 | 31 (16%) | 194 | 36.3 | 52 (27%) | ***0.95 (0.76,1.17)*** | ***-0.107 (-0.193, -0.02)*** |

When the HAI GMTs were summarized separately for the two study years, we observed higher values in the 2018-2019 season than 2019-2020 across all strains and for both aTIV and HD-IIV3 vaccines (Supplemental Table 2). The rates of HAI seroconversion were calculated for each individual season and generally show a similar pattern of responses, with the highest seroconversion rates observed for H3N2 strain in both seasons and both vaccines (Supplemental Table 3).

The proportion of participants who achieved seroprotection, defined as an HAI titer ≥40 at day 28, was similar between the aTIV and HD-IIV3 groups regardless of strain and observed at higher rates in 2018-2019 than in 2019-2020 across strains and vaccines (Supplemental Table 4). In both vaccines, over 80% seroprotection was observed for all strains when considering the seasons combined. In a subset of 2018-2019 season subjects at day 180 we measured HAI titers and observed a decrease from day 28 in both vaccine groups (Supplemental Figure 1).

### Secondary Outcomes

When the non-inferiority criteria were applied to neuraminidase inhibition (NI) titers, aTIV was non-inferior to HD-IIV3 for both seroconversion and GMT at day 28 post-vaccination for both H1N1 and H3N2 strains. For H1N1, the NA seroconversion rate was 62% for aTIV compared to 29% for HD-IIV3, and for H3N2, the rates were 27% and 16%, respectively. NI GMT values for H1N1 were 351.5 for aTIV and 159.4 for HD-IIV3, while for H3N2, the NI GMTs were 36.3 and 34.4, respectively (Table 3).

In a subset of 2018-2019 season subjects at day 180 we measured NI titers and observed a decrease from day 28 in both vaccine groups for H1N1 with little to no decline observed for H3N2 (Supplemental Figure 1).

In the 2019-2020 season, we summarized HAI and NI for the participants who received RIV (n=32). Compared to aTIV and HD-IIV3 in the same season, RIV produced similar HAI titers. Unlike the aTIV and HD-IIV3 vaccines which contain some neuraminidase, RIV does not; subjects administered RIV had less change in NI from day 0 to Day 28 than those who received the other vaccines (Supplemental Figure 2).

## Discussion

The US Advisory Committee on Immunization Practices (ACIP) recommends that individuals aged 65 and older receive an enhanced vaccine that includes two high-dose vaccines (HD-IIV3 and RIV) and adjuvanted influenza vaccine (aTIV) ^2^. Each of these enhanced vaccines has demonstrated better clinical efficacy or effectiveness than standard-dose vaccines ^3, 4, 6, 10^, and greater immunogenicity^11^. Three studies have directly compared the enhanced vaccines in older individuals and none in long-term care residents ^12-14^. This study provides the opportunity to contrast immunogenicity data with these 3 clinical effectiveness studies although our subjects are all long-term residents and the effectiveness studies are a general population of older individuals. Each of these effectiveness studies use metadata from large database systems with the strengths and weaknesses of such designs. Between the studies, they span 4 influenza seasons with two studies overlapping in one season. They all focus on the reduction of influenza associated clinical outcomes assessed primarily by inpatient or outpatient healthcare visits. The Imran et al and Boikes et al each overlap one year of our study.

Our immunogenicity studies observed a number of differences between the vaccines, strains and seasons. Our results demonstrate that aTIV failed to meet non-inferiority criteria for HAI seroconversion at day 28 post-vaccination for all three influenza strains when both flu seasons were combined. The small percentage differences, although statistically significant, may not represent a clinically important finding, considering the seroconversion rates for aTIV were 39.6% for H1N1, 54.1% for H3N2, and 35.6% for the B strain, compared to 43.7%, 64.2%, and 43.0%, respectively, for HD-IIV3. Despite the differences in HAI seroconversion rates, aTIV was non-inferior to HD-IIV3 in terms of NI titers. At day 28 post-vaccination, aTIV demonstrated higher seroconversion rates and NI GMTs for H1N1 and H3N2 compared to HD-IIV3.

The immunogenicity results favored HD-IIV3 in HAI titer seroconversion but favored aTIV in anti-NA titers. These distinct and opposing advantages may help explain the somewhat contradictory reported clinical effectiveness of which vaccine is better. Two studies found aTIV more effective relative to HD-IIV3 but in the modest range of a 7% reduction in influenza-related medical encounters (IRME) in those over age 65 and in a different season a 10-18% reduction in IRME specifically older persons CDC defined influenza risk factors and no difference in the older population without influenzas risk factors ^13, 14^. In contrast, Van Aalst et al found that HD-IIV3 was 12 % better than aTIV at reducing respiratory hospital admissions across two H3N2 predominant seasons ^12^.

The current study adds to the prior data by demonstrating potential relevance of anti-NA titers to protection. Several prior studies also indicate the importance and relevance of anti-NA titer to clinical effectiveness ^15, 16^. Yet, RIV does not have any NA in it but nonetheless has proven as a highly effective vaccine among non-institutionalized older adults ^10^. Thus we expect the role for anti-NA as a complementary one, and potentially independent from the protection provided by anti-HA antibody.

Our small group of nursing home residents who received RIV produced HAI titers similar to those of residents who received either of the other vaccines. Therefore, purely from an immunogenicity standpoint, this supports the inclusion of RIV in the enhanced category as assigned by the ACIP for preferential recommendation for older adults. Dunkle et al compared RIV to standard dose vaccine in those age 50 and older and found RIV to be superior to standard dose vaccine ^10^.

### Limitations

While we utilized standard assays to assess humoral immunity, antibody responses in older long-term care residents have less consistent reliability as indicators of protection. Cellular immunity may play a more significant role in determining vaccine effectiveness in this population, and this was not assessed in our study. NI and HAI responses may also provide a proxy for cellular immunity, and evidence for protection that could supplant or eclipse the benefit provided by just increased antibody titers. While antibody titers can serve to prevent or reduce the impact of the initial infection, cellular immunity associates with clinical recovery, and therefore remains an unmeasured confounder of this analysis. Also, we did not measure antibody avidity, which may differ for these vaccines ^11^. Finally, participants with conditions that could impair immune response such as the use of immunomodulatory drugs or recent cancer requiring chemotherapy were excluded from the study, potentially limiting the generalizability of our findings to those individuals. Future studies should explore vaccine efficacy in this highly frail subset of the population.

### Conclusion

Both aTIV and HD-IIV3 elicited strong humoral immune responses in older individuals residing in long-term care facilities. While aTIV did not meet non-inferiority standards for HAI seroconversion, it demonstrated non-inferiority for neuraminidase inhibition and outperformed HD-IIV3 in terms of NA seroconversion and GMTs. On balance, our data support the clinical and immunogenicity data from others and add nuance with respect to NI; our results support the ACIP recommendations for offering an enhanced to older individuals.

## Data Availability

All data produced in the present study are available upon reasonable request to the authors after publication in a peer-reviewed journal

## Notes

### Author contributions

E.D and J.K. performed investigations, data organization, draft preparation. B.W. performed statistical analyses and manuscript review and editing. D.W. and S.F carried out investigations and data. D.K., E.B., S.R. and R.B. were responsible for project administration. D. C. and S. G. were responsible for project conceptualization, methodology, project administration, supervision, and manuscript review and editing.

### Disclaimer

The contents do not represent the views of the U.S. Department of Veterans Affairs or of the United States Government.

## Acknowledgments

We would like to thank the long-term care facility staff and residents who participated in this study, without whom this study would not have been feasible. The sponsor (NIH) plaed no roll in this study other than funding fuding it.

## Supplementary Data

**Table S1.** Statistical summary of D0 standardized mean difference in outcomes by year and combined. Values below |0.1| were considered well-balanced by randomization, and values exceeding |0.25| were considered strongly unbalanced.

| Assay | Strain | 2018-2019 | 2019-2020 | Combined |
| --- | --- | --- | --- | --- |
| NI | H1N1 | -0.19 | 0.11 | -0.02 |
| NI | H3N2 | 0.01 | -0.11 | -0.08 |
| HAI | B | -0.13 | 0.08 | -0.01 |
| HAI | H1N1 | 0.12 | -0.08 | 0.05 |
| HAI | H3N2 | 0.12 | 0.03 | 0.09 |

**Table S2.** Statistical summary of GMT values for all assays at D0 and D28.

|  |  |  | HD-IIV3 |  |  |  | aTIV |  |  |  |
| --- | --- | --- | --- | --- | --- | --- | --- | --- | --- | --- |
|  |  |  | 2018-2019 |  | 2019-2020 |  | 2018-2019 |  | 2019-2020 |  |
| Assay | Strain | day | N | GMT (95% CI) | N | GMT (95% CI) | N | GMT (95% CI) | N | GMT (95% CI) |
| NI | H1N1 | 0 | 83 | 51.4<br>(39,67.7) | 106 | 108.1<br>(77.8,150.2) | 89 | 40.9<br>(32.2,52) | 103 | 129<br>(94.3,176.4) |
| NI | H1N1 | 28 | 83 | 71.8<br>(53,97.3) | 106 | 297.8<br>(214.3,413.8) | 89 | 167.7<br>(128.2,219.3) | 103 | 666.4<br>(497.7,892.3) |
| NI | H3N2 | 0 | 86 | 15.3<br>(13.2,17.8) | 106 | 26.7<br>(21.9,32.5) | 91 | 15.4<br>(13.7,17.5) | 103 | 23.8<br>(19.6,28.9) |
| NI | H3N2 | 28 | 86 | 24.9<br>(20.6,30.1) | 106 | 44.7<br>(35.4,56.4) | 91 | 27.3<br>(23.3,32.1) | 103 | 46.7<br>(37.1,58.8) |
| HAI | B | 0 | 86 | 56.8<br>(41.2,78.3) | 107 | 27<br>(21.2,34.2) | 91 | 47.5<br>(37.4,60.3) | 103 | 29.8<br>(23.4,38.1) |
| HAI | B | 28 | 86 | 210.4<br>(152.5,290.3) | 107 | 66.1<br>(51.1,85.5) | 91 | 113.6<br>(87.9,146.8) | 103 | 75.5<br>(58.1,98.1) |
| HAI | H1N1 | 0 | 83 | 73<br>(52.7,101) | 107 | 22.4<br>(18,27.8) | 89 | 87.8<br>(63.6,121.3) | 103 | 20.5<br>(16.7,25.2) |
| HAI | H1N1 | 28 | 83 | 261.9<br>(192.3,356.6) | 107 | 67.2<br>(52.3,86.3) | 89 | 213.4<br>(156.9,290.4) | 103 | 70.5<br>(53.5,92.9) |
| HAI | H3N2 | 0 | 86 | 119.7<br>(89.1,160.8) | 107 | 16.9<br>(14.2,20) | 91 | 141.6<br>(103.4,194) | 103 | 17.3<br>(14.4,20.7) |
| HAI | H3N2 | 28 | 86 | 517.3<br>(380.8,702.6) | 107 | 120.4<br>(87.2,166.2) | 91 | 375.5<br>(271.3,519.7) | 103 | 140.2<br>(96.9,202.7) |

**Table S3.** Statistical summary of HAI and NI seroconversion rates by year.

| Assay | Strain | HD-IIV3 |  |  |  | aTIV |  |  |  |
| --- | --- | --- | --- | --- | --- | --- | --- | --- | --- |
|  |  | 2018-2019 |  | 2019-2020 |  | 2018-2019 |  | 2019-2020 |  |
|  |  | N | % Seroconversion (95% CI) | N | % Seroconversion (95% CI) | N | % Seroconversion (95% CI) | N | % Seroconversion (95% CI) |
| NI | H1N1 | 83 | 9.6 (4.6,18.6) | 106 | 43.4 (33.9,53.4) | 89 | 64 (53.1,73.7) | 103 | 60.2 (50.1,69.6) |
| NI | H3N2 | 86 | 10.5 (5.2,19.4) | 106 | 20.8 (13.7,29.9) | 91 | 20.9 (13.3,30.9) | 103 | 32 (23.4,42.1) |
| HAI | B | 86 | 47.7 (36.9,58.7) | 107 | 39.3 (30.1,49.2) | 91 | 34.1 (24.7,44.8) | 103 | 36.9 (27.8,47) |
| HAI | H1N1 | 83 | 44.6 (33.8,55.9) | 107 | 43 (33.6,52.9) | 89 | 29.2 (20.3,40) | 103 | 48.5 (38.7,58.5) |
| HAI | H3N2 | 86 | 57 (45.9,67.5) | 107 | 70.1 (60.4,78.4) | 91 | 35.2 (25.6,46) | 103 | 70.9 (61,79.2) |

**Table S4.**
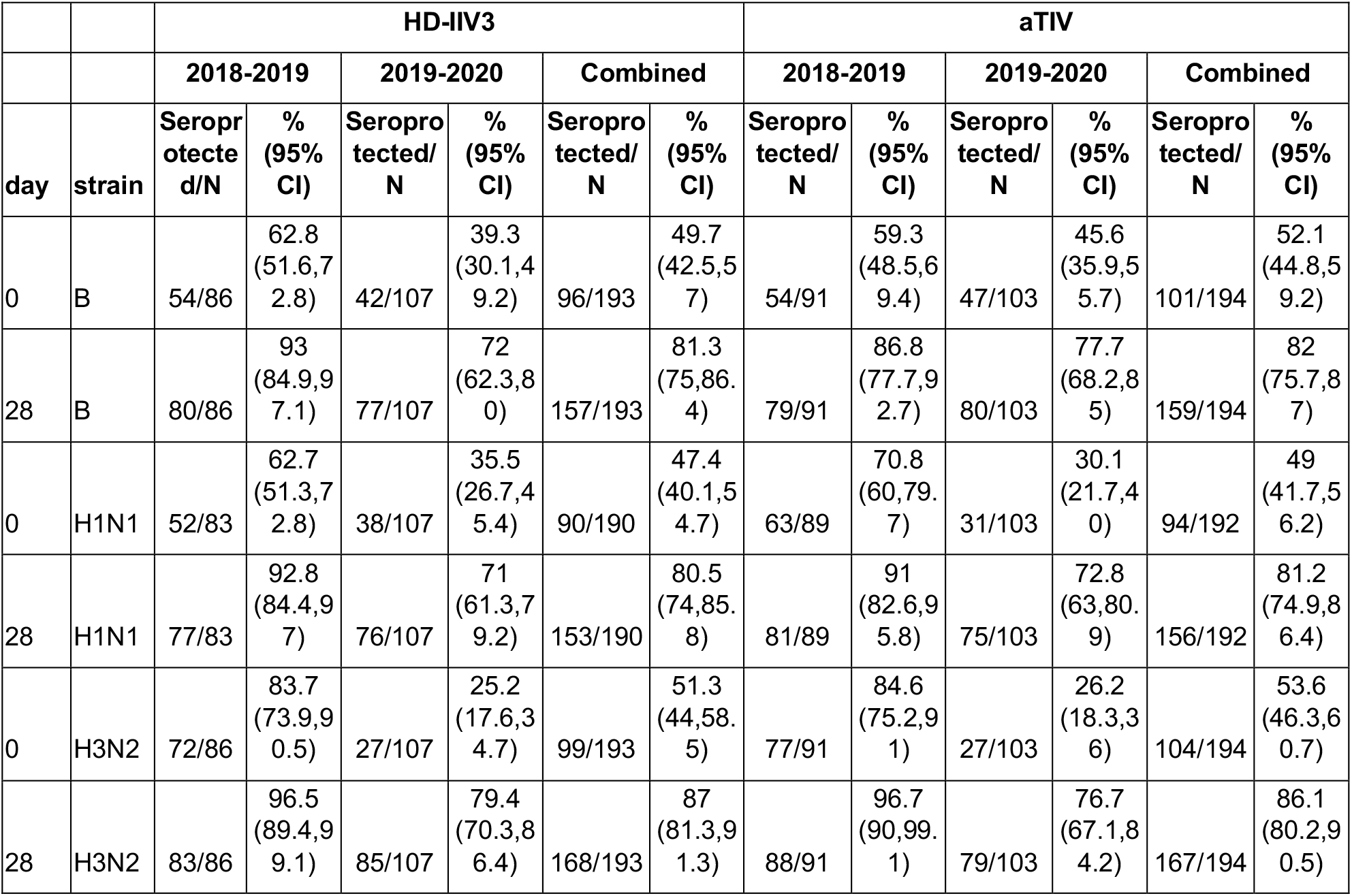
Statistical summary of HAI seroprotection rates by year and combined.

**Supplemental Figure 1.**
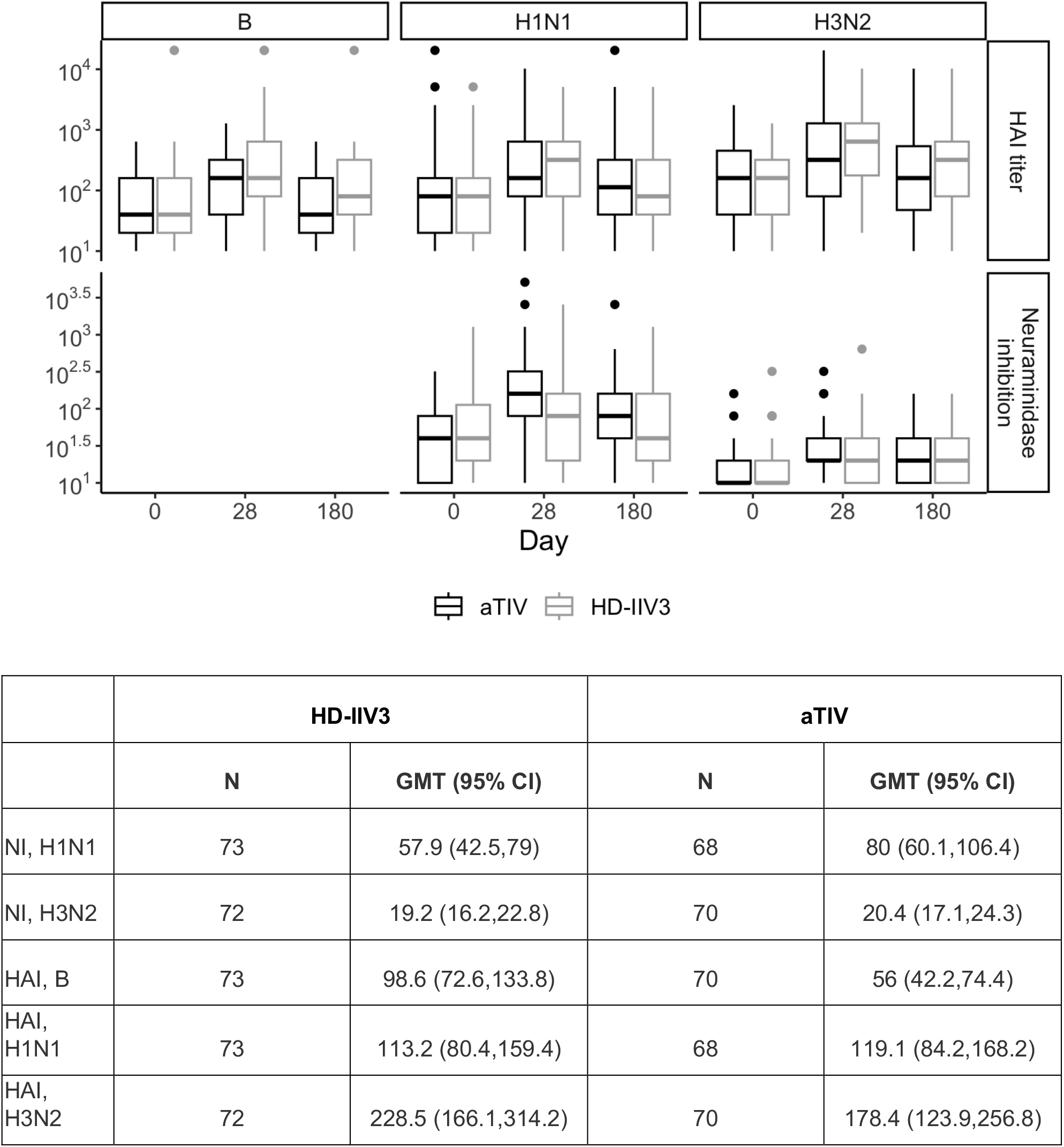
HAI and NI GMT titers at D180 for year 1 in figure and table format.

**Supplemental Figure 2.**
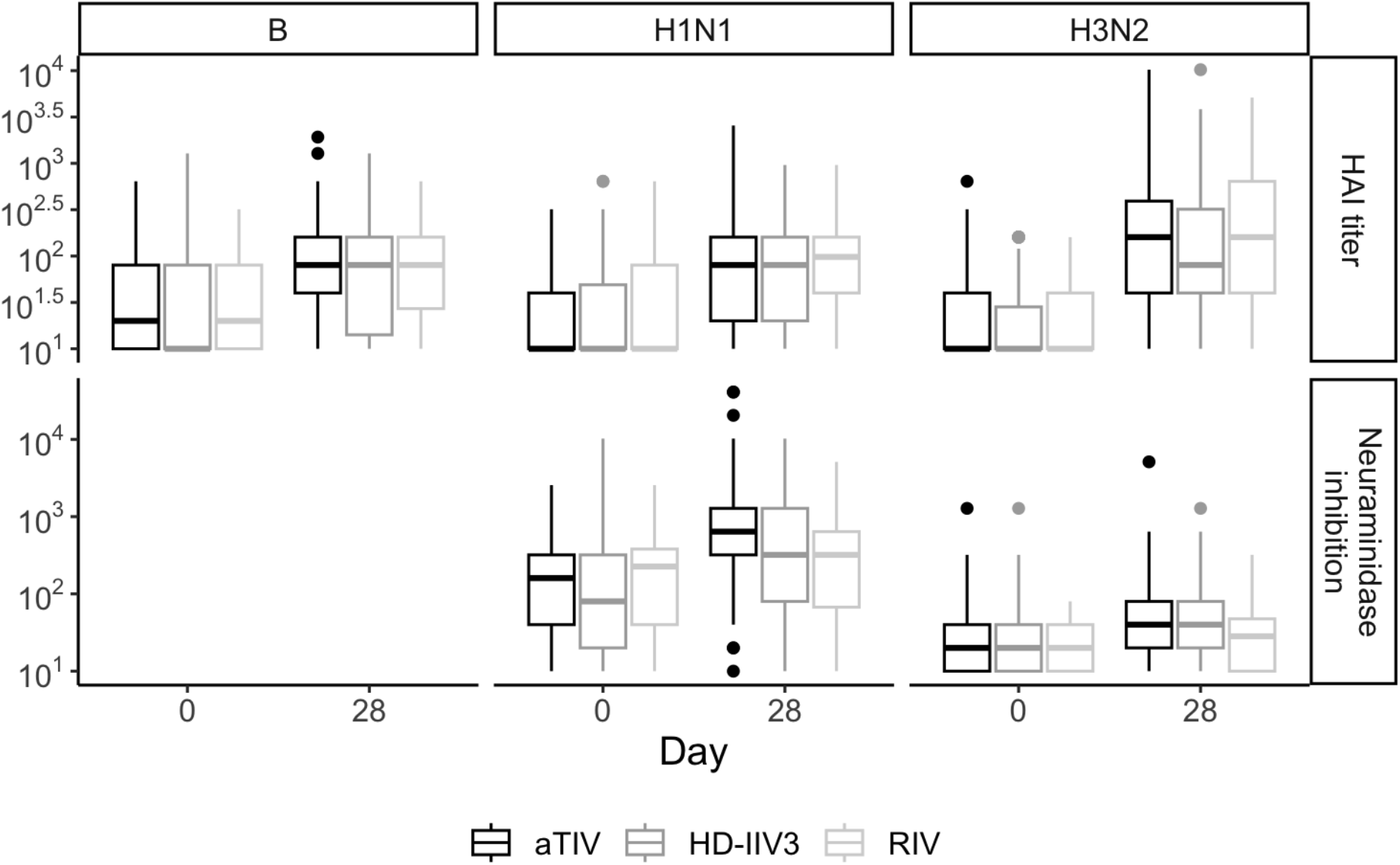
Observed distributions of HAI and NI titers for aTIV, HD-IIV3, and RIV, by day and strain for 2019-2020 season.

